# PERFORMANCE AND UTILITY OF AN ORAL FLUID-BASED RAPID POINT-OF-CARE TEST FOR SARS-COV-2 ANTIBODY RESPONSE FOLLOWING COVID-19 INFECTION OR VACCINATION

**DOI:** 10.1101/2021.06.28.21259657

**Authors:** Paturi V. Rao, Dhanalakshmi Nair-Shaef, Siting Chen, Steven C. Kazmierczak, Charles T. Roberts, Srinivasa R. Nagalla

**Affiliations:** Diabetomics, Inc., Hillsboro, OR 97006; COVYDx, Inc., Hillsboro, OR 97006; Oregon Health & Science University, Portland OR 97239

## Abstract

Analysis of anti-SARS-CoV-2 antibodies can identify recent-onset or prior COVID-19 infection or vaccine-induced humoral immunity. We have developed a rapid point-of-care test for IgG, M, or A-class immunoglobulins that recognize the S1 domain of the SARS-CoV-2 spike protein (CovAb™). The test employs a lateral-flow strip design with a recombinant SARS-CoV-2 spike protein S1 domain capture antigen to detect anti-SARS-CoV-2 antibodies in oral fluid samples. Oral fluid samples are collected with a swab that captures the gingival crevicular fluid component of oral fluid that represents a plasma transudate and that is the primary source of oral fluid monomeric antibodies. The sensitivity of the CovAb™ test is 97.29% and the specificity is 98.13%, and the results obtained are similar to those obtained using matched fingerstick whole blood samples and in an EUA-approved commercial serology test. Oral fluid SARS-CoV-2 antibodies could be detected in subjects more than 7 months post-symptom onset. We also demonstrate the utility of the CovAb™ test in characterizing adaptive immune responses to vaccination in COVID-19-naïve and exposed populations after first and second vaccine doses and show that significant heterogeneity in magnitude of antibody titers achieved is seen after both doses and that prior COVID-19 exposure increases the adaptive immune response to vaccination.

## INTRODUCTION

The COVID-19 pandemic caused by the novel SARS-CoV-2 coronavirus (1) has created a worldwide need for effective diagnostics to enable identification and contact tracing of infected individuals to administer appropriate clinical care as necessary and to contain further disease spread, as well as to assess serological responses to infection and vaccines (2-4). PCR-based methods for detection of viral nucleic acid are sensitive but require centralized laboratory facilities, an appreciable turnaround time, and do not reliably identify disease experienced more that 14 days before testing nor the presence of neutralizing antibodies that may indicate immunity. Antigen tests that detect SARS-CoV-2 viral proteins are more amenable to rapid, point-of-care designs, but are less sensitive than PCR tests and also only identify ongoing or recent disease and do not reveal the presence of COVID-19 antibodies.

Tests specifically designed to detect anti-SARS-CoV-2 antibodies can provide information on the adaptive immune response to recent disease or vaccination, but test performance can be variable (5,6), although some tests exhibit high sensitivity and specificity (7-10). There are a number of possible test designs that influence the type of information obtained. Major variables include the capture antigen (nucleocapsid vs spike protein S1 and/or S2 domains vs receptor-binding domain), the immunoglobulin class detected (IgG, M, or A), the clinical sample source (blood, serum, plasma, or oral fluid), and the analytical design (ELISA/chemiluminescent immunoassay, lateral-flow, etc.). A spike protein receptor-binding domain-based test would be more likely to detect potential neutralizing antibodies as they would interfere with viral attachment to target cells (11). In support of this concept, the most potent neutralizing antibodies isolated from convalescent COVID-19 patients target the SARS-CoV-2 spike protein, particularly the receptor-binding domain (12-14). Additionally, the S1 domain (which includes the receptor-binding domain) is less conserved that the S2 domain or nucleocapsid between seasonal SARS and SARS-CoV-2, thus making the use of the S1 or receptor-binding domain for antibody capture potentially more specific for COVID-19 (15). The various detection platforms include standard PCR instruments for nucleic acid detection, plate or automated analyzer platforms for ELISA/chemiluminescent assays, and lateral-flow devices for some rapid-detection tests.

Several lateral-flow COVID-19 serology tests have been developed that employ fingerstick whole blood samples and that exhibit good performance (16), including in a point-of-care (POC) setting (17). While the majority of current tests utilize a nasopharyngeal swab for PCR testing or a blood sample (whole blood, serum, or plasma) for antigen and antibody testing, saliva has emerged as a noninvasive alternative for SARS-CoV-2 nucleic acid (18) and antigen (19,20) detection based on SARS-CoV-2 infection of the oral cavity (21) enabled by oral cavity expression of SARS-CoV-2 entry molecules (22). Saliva has also been reported to be a source of SARS-CoV-2 antibodies (23-26). Our approach was to utilize the optimal combination to produce a simple, sensitive, cost-effective, rapid POC design for detection of anti-SARS-CoV-2 antibodies in oral fluid, specifically the gingival crevicular fluid (GCF) that is a transudate of plasma (27-28), and which is increasingly appreciated as an important source of diagnostic biomarkers (29-33). The use of GCF as the assay fluid enables a completely non-invasive format. This test (CovAb™) employs a recombinant SARS-CoV-2 spike protein S1 domain as a capture antigen and a lateral-flow strip that detects IgG, A, and M immunoglobulins, since all three immunoglobulin classes are components of the SARS-CoV-2-induced adaptive immune response (34,35).

## METHODS AND RESULTS

### CovAb™ test design

Oral fluid SARS-CoV-2 antibodies were assayed using the CovAb™ SARS-CoV-2 Ab test (FDA EUA authorization #210017 dated 6/4/2021), a lateral-flow immunochromatography assay. Recombinant SARS-Cov-2 spike protein S1 domain produced in human embryonic kidney HEK293 cells (Diabetomics, Inc.) and goat anti-human IgG, IgA, and IgM (Jackson ImmunoResearch Laboratories, Inc.) were used for capture and detection of SARS-CoV-2 antibodies. For quantification of antibody levels shown in Tables 8 and 9 and Figures 1-3, a proprietary lateral-flow quantification reader based on image capture technology (Diabetomics, Inc.) was used. Samples that registered above 70 reflectance units with the reader were considered positive based on the maximal level observed in 160 SARS-CoV-2 RT-PCR-negative GCF samples.

**Figure 1.**
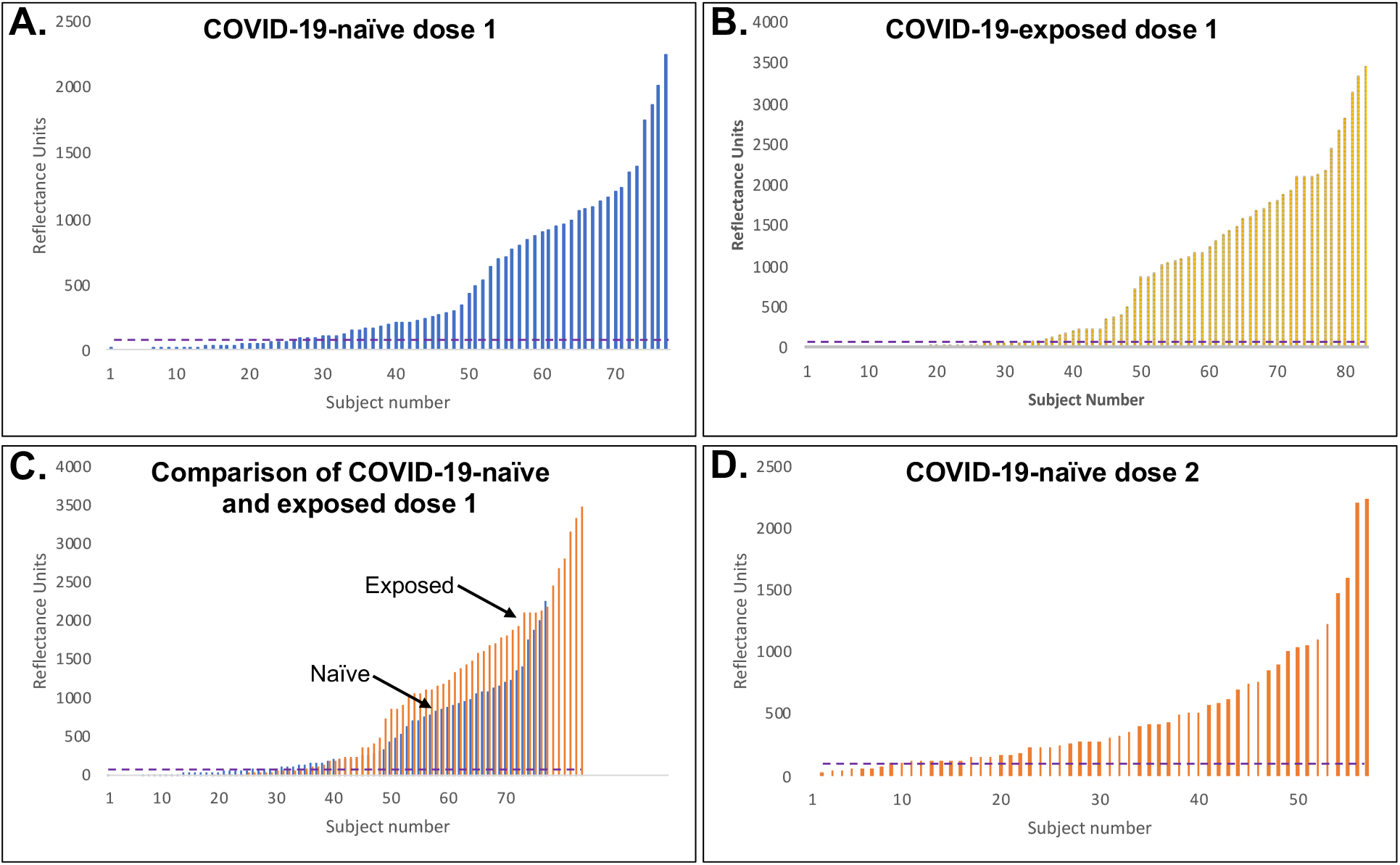
Distribution of antibody responses to vaccination. A. Distribution of CovAb™ test values in 77 COVID-19-naïve study subjects after first vaccine dose. B. Distribution of CovAb™ test values in 83 COVID-19-exposed study subjects after first vaccine dose. C. Comparison of CovAb™ test values in COVID-19-naïve and exposed study populations. D. Distribution of CovAb™ test values in 57 COVID-19-naïve study subjects after second vaccine dose. The horizontal dotted line in each graph represents the 70 reflectance-unit threshold for a positive result based on the maximal level observed in 160 SARS-CoV-2 RT-PCR-negative GCF samples.

### Cross-reactivity/analytical specificity studies

#### Cross-reactivity with virus, bacteria, and auto-antibodies

Serum or plasma samples validated for the presence of antibodies to specific pathogens (BioIVT, NY) were spiked into negative GCF samples and tested for cross-reactivity with the CovAbTM test. The pathogens tested included influenza A, influenza B, Haemophilus influenza, respiratory syncytial virus (RSV), hepatitis C virus (HCV), hepatitis B virus (HBV), human immunodeficiency virus (HIV), alpha coronavirus 229E, alpha coronavirus NL63, beta coronavirus OC43, beta coronavirus HKU1, enterovirus, M. pneumoniae, C. pneumoniae, B. pertussis, Parainfluenza 1-4, and rheumatoid factor (RF). As shown in **Table 1**, there was no cross-reactivity of any of the samples with the CovAb™ test.

**Table 1.** Cross-reactivity with virus, bacteria, and auto-antibodies.

| Table 1. Cross-reactivity with virus, bacteria, and auto-antibodies. |  |  |  |  |
| --- | --- | --- | --- | --- |
| Virus/bacteria/autoantibody-positive sample | Source | Sample type for spiking | Number reactive | Number non-reactive |
| Influenza A | BioIVT,<br>Hicksville, NY<br>www.bioivt.com | Plasma | 0 | 5 |
| Influenza B |  | Plasma | 0 | 7 |
| H. influenza |  | Plasma | 0 | 8 |
| Respiratory syncytial virus (RSV) |  | Plasma | 0 | 9 |
| Hepatitis C virus (HCV) |  | Serum | 0 | 5 |
| Hepatitis B virus (HBV) |  | Serum | 0 | 5 |
| Human immunodeficiency virus (HIV) |  | Serum | 0 | 5 |
| Alpha coronavirus 229E |  | Serum | 0 | 20 |
| Alpha coronavirus NL63 |  | Serum | 0 | 17 |
| Beta coronavirus OC43 |  | Serum | 0 | 17 |
| Beta coronavirus HKU1 |  | Serum | 0 | 12 |
| Enterovirus |  | Plasma | 0 | 4 |
| M. pneumoniae |  | Plasma | 0 | 11 |
| C. pneumoniae |  | Plasma | 0 | 3 |
| B. pertussis |  | Plasma | 0 | 5 |
| Parainfluenza 1-4 |  | Plasma | 0 | 12 |
| Rheumatoid factor (RF) | Diabetomics, Inc. | Serum | 0 | 5 |

#### Interference by exogenous substances

The CovAb™ test uses an oral fluid specimen (primarily GCF). Robustness testing included the testing of exogenous substances that may be expected to be consumed orally. Substances tested included ethanol, nicotine, hot caffeinated beverages (coffee and tea), carbonated sodas (Coca-Cola), orange juice, mouthwash (Listerine), and cough syrup (Nyquil Cough DM+, spiked at 7% total volume). In brief, stocks of nicotine, ethanol, and cough syrup were made and diluted in pooled GCF to obtain testing matrices containing these interferents. For the other potential interferents, volunteers used or consumed the potential interferents prior to sampling. Each matrix was then tested with 5 negatives, 5 low positives, and one high positive. GCF samples were collected from six different individuals before consuming the interferent, immediately after consuming the interferent, and 30 minutes after consuming the interferent (total of 3 testing points). None of these substances affected the development of the control or test lines (data not shown).

### Clinical agreement studies using oral fluid

In a prospective clinical study, 306 subjects were enrolled at 4 study sites (Oregon Health & Science University, Portland, OR; two institutional hospitals in Hyderabad, Telangana, India, dedicated for COVID-19 care, the Asian Institute of Gastroenterology (AIG) and Sunshine specialty hospitals (SUNSEC); and at two nursing homes in Seattle, WA) to collect GCF samples for testing. Samples were obtained at early (2-7 days from onset of symptoms), intermediate (8-14 days), and convalescent (>14 days) stages of disease. 146 subjects were SARS-CoV-2 RT-PCR-positive and 160 subjects were SARS-CoV-2 RT-PCR-negative based on CDC emergency use authorization (EUA)-approved RT-PCR tests. The demographics of the study population are shown in **Table 2**. The positive percent agreements (PPA) and negative percent agreements (NPA) are shown in **Table 3**, with the positive cases distributed by days from symptom onset. This study showed that, in the early-onset stage, antibody levels are low and variable, and increase at the intermediate stage with a higher percent positivity. In the convalescent stage, seroconversion is complete and a high percentage of subjects developed detectable levels of antibodies. This study demonstrated 97.29% PPA agreement in seroconverted subjects (>14 days from onset of symptoms) and 98.13% NPA.

**Table 2.** Demographics of the test performance study population.

| Table 2. Demographics of the test performance study population. |  |
| --- | --- |
| Age (years) |  |
| 14-24 | 15 |
| 25-50 | 123 |
| 51-64 | 58 |
| 65-86 | 50 |
| Not specified | 60 |
| Gender |  |
| Female | 112 |
| Male | 156 |
| Not specified | 38 |
| Ethnicity |  |
| African-American | 36 |
| Asian | 9 |
| Caucasian | 108 |
| Filipino | 10 |
| Hispanic | 17 |
| Southeast Asian | 126 |

**Table 3.**
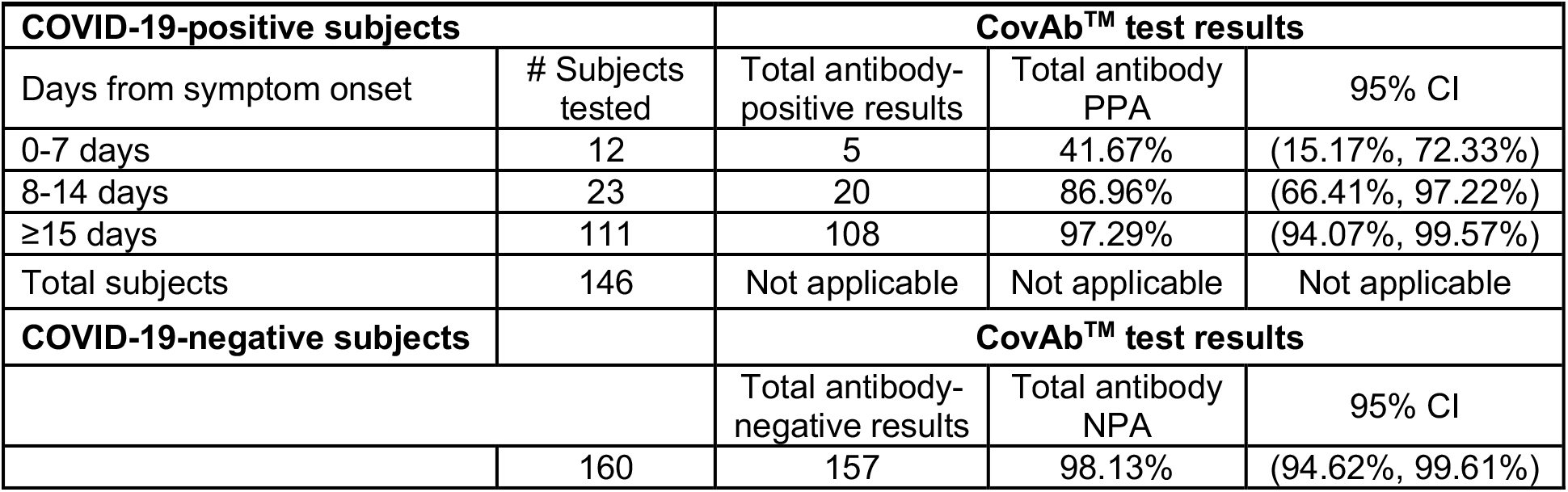
CovAb™ test positive percent agreement (PPA) and negative percent agreement (NPA).

We also determined the persistence of oral fluid antibodies in the antibody-positive subjects. As shown in **Table 4**, the CovAb™ test detected antibodies up to 7.2 months post-symptom onset in >8% of the antibody-positive group.

**Table 4.** Persistence of antibody response in the COVID-19-naive group.

| Table 4. Persistence of antibody response in the COVID-19-naive group. |  |  |
| --- | --- | --- |
| Days from symptom onset | Number of subjects positive in CovAb™ test | % seropositive |
| 3-30 days | 39 | 26.7 |
| 31-60 days | 28 | 19.2 |
| 61-90 days | 39 | 26.7 |
| 91-120 days | 14 | 9.6 |
| 121-150 days | 5 | 3.4 |
| 151-180 days | 9 | 6.2 |
| 181-218 days | 12 | 8.2 |
| **Total** | **146** | **100** |

### Comparison of oral fluid with fingerstick sample and with RT-PCR test status

A subset of the subjects (n=83) from the US nursing home study population provided fingerstick whole blood samples. This study set included 54 subjects who tested positive for SARS-CoV-2 and 29 subjects who tested negative for SARS-CoV-2 by an RT-PCR test. As shown in **Table 5**, there was 98% PPA between the CovAb™ fingerstick test and RT-PCR-positive status and 100% PPA between the CovAb™ fingerstick and GCF samples.

**Table 5.** Agreement between RT-PCR test status and CovAb™ test fingerstick whole blood and GCF test results.

|  |  |  |  |
| --- | --- | --- | --- |
| SARS-CoV-2 RT-PCR test result | CovAb™ fingerstick test result |  |  |
|  | - | + | Total |
| - | 28 | 1 | 29 |
| + | 1 | 53 | 54 |
| Total | 29 | 54 | 83 |
| **Positive Percent Agreement (PPA) $100 \times 53/54 = 98.14\%$** |  |  |  |
| **Negative Percent Agreement (NPA) $100 \times 28/29 = 96.55\%$** |  |  |  |
| CovAb™ fingerstick test result | CovAb™ GCF test result |  |  |
|  | - | + | Total |
| - | 28 | 1 | 29 |
| + | 0 | 54 | 54 |
| Total | 29 | 54 | 83 |
| **Positive Percent Agreement (PPA) $100 \times 54/54 = 100\%$** |  |  |  |
| **Negative Percent Agreement (NPA) $100 \times 28/29 = 96.55\%$** |  |  |  |

### Comparator EUA serology test (Siemens total antibody test)

Fingerstick whole-blood samples were obtained from 38 subjects who tested positive with the CovAb™ test using GCF, and all were also positive in the CovAb™ test using these whole blood samples. 36 of the fingerstick samples had sufficient volume to also be tested with an US FDA EUA-authorized serology test (Siemens total antibody test). This test uses the same spike protein S1 domain for capture as the CovAb™ test, and exhibits high specificity and sensitivity in head-to-head comparisons with other commercial serology tests (7). As shown in **Table 6**, there was excellent agreement between the two tests for 35/36 samples, with 1 sample that was RT-PCR-positive and positive in the CovAb™ test being negative in the Siemens test (CovAb™ test PPA, 100%; Siemens test PPA >97%).

**Table 6.** Comparison of CovAbTM and Siemens EUA serology tests.

| **Table 6. Comparison of CovAbTM and Siemens EUA serology tests.** |  |  |
| --- | --- | --- |
| **Comparator EUA test result** | **Negative** | **Positive** |
| CovAbTM test fingerstick-positive | 1 | 35 |
| CovAbTM test fingerstick-negative | 0 | 0 |
| Total | 1 | 35 |

### Assessment of post-vaccination antibody responses

A prospective study of 160 subjects was conducted to examine post-vaccination antibody responses at Ramadev Rao Hospital, Telengana, Hyderbad, India. 77 COVID-19-naïve subjects who tested negative with the CovAb*™* test prior to screening were tested at 28-29 days after the first dose of the Oxford-AstraZeneca vaccine (Covishield). 57 subjects from the same cohort were tested 14 days after the second dose of the vaccine. 83 subjects who were exposed to COVID-19 (clinical laboratory staff performing COVID-19 testing at Tennet Diagnostics, Hyderabad, India; the COVID-19-exposed group) was tested at 18-20 days after the first dose of the vaccine. The demographics of these groups are shown in **Table 7**. The median, 25^th^, and 75^th^ percentile response values, and the interquartile range (IQR) for the positive samples in the three groups (with reflectance unit readings >70) are shown in **Table 8**.

**Table 7.** Demographics of the antibody response study population.

| **Table 7.**<br>**Demographics of the antibody response study population.** |  |
| --- | --- |
| **Age (years)** |  |
| 14-24 | 59 |
| 25-50 | 75 |
| 51-64 | 19 |
| 65-86 | 7 |
| **Gender** |  |
| Female | 62 |
| Male | 98 |
| **Ethnicity** |  |
| Southeast Asian | 160 |

**Table 8.** Parameters of post-vaccine antibody responses.

| **Table 8. Parameters of post-vaccine antibody responses.** |  |  |  |  |  |  |  |
| --- | --- | --- | --- | --- | --- | --- | --- |
| **Group** | **n** | **Negative (%)** | **Positive (%)** | **Median\*** | **25<sup>th</sup> percentile\*** | **75<sup>th</sup> percentile\*** | **IQR\*** |
| **COVID-19-naïve (dose 1)** | 77 | 24 (33) | 53 (67) | 488 | 178 | 990 | 812 |
| **COVID-19-exposed (dose 1)** | 83 | 32 (37) | 51 (63) | 1163 | 360 | 1879 | 1519 |
| **COVID-19-naïve (dose 2)** | 57 | 7 (14) | 50 (86) | 335.5 | 167 | 748 | 581 |

\*Values for positive samples only.

**Table 9.**
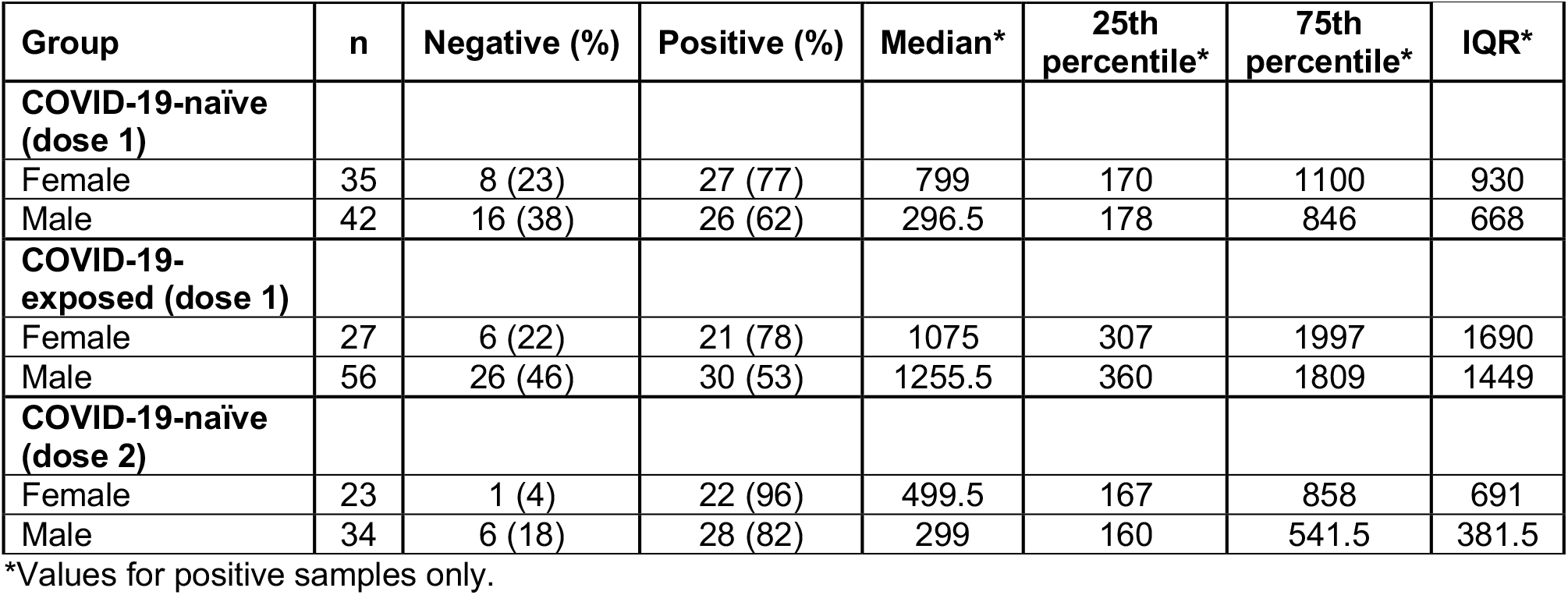
Parameters of post-vaccine antibody responses in females and males.

The response of the COVID-19-naïve group to the first vaccine dose is shown in **Figure 1A**. Of this group, 24 (33%) were antibody-negative and 53 (67%) were antibody-positive, with a median response of 488 reflectance units (**Table 8**). The response of the COVID-19-exposed group to the first vaccine dose is shown in **Figure 1B**. Of this group, 32 (37%) were antibody-negative and 51 (63%) were antibody-positive, a proportion similar to the COVID-19-naïve group. However, as illustrated by the comparison of the responses of the COVID-19-naïve and exposed groups shown in **Figure 1C**, the response of the COVID-19-exposed group was more robust, with a median response of 1163 reflectance units, and more evenly distributed, with a larger IQR (**Table 8**), and as illustrated in **Figure 2**, the difference in the median response in antibody-positive COVID-naïve subjects was significantly greater that that seen in the COVID-19-naïve group (p=0.0005).

**Figure 2.**
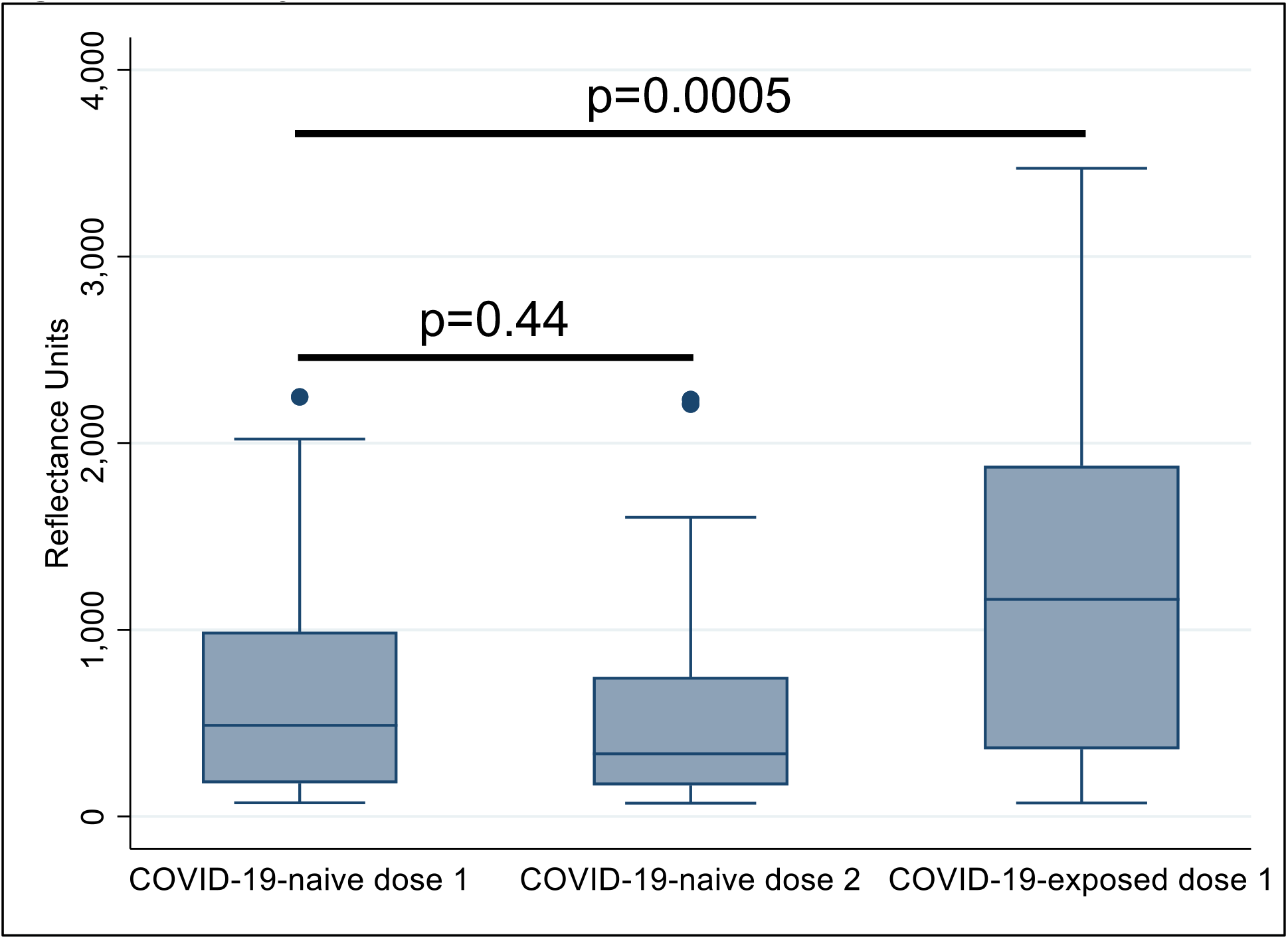
Antibody responses to vaccination. P values are based on a two-sample Wilcoxon rank-sum (Mann-Whitney) test.

The response of the COVID-19-naïve group to a second vaccine dose is shown in **Figure 1D**. In this group, 7 (14%) were antibody-negative and 50 (86%) were antibody-positive, representing a higher rate of antibody response vs the first dose (86 vs 67%). However, as also illustrated in **Figure 2**, there was no statistically significant difference between the median antibody responses of the COVID-naïve group to the first and second doses of vaccine (p=0.44), although this may reflect the shorter post-vaccination interval for the second dose (i.e., 14 vs 28-29 days).

We also investigated the effect of sex on vaccine responses. As shown in **Table 9**, the rate of positive responses in all three groups was greater for females that males (77 vs 62%, 78 vs 53%, and 96 vs 82%, respectively, in the COVID-19-naïve dose 1, COVID-exposed dose 1, and COVID-19-naïve dose 2 groups. The median response was also greater in females than males in the COVID-19-naïve dose 1 and COVID-19-naïve dose 2 groups (799 vs 296.5 and 499.5 vs 299, respectively), while the median response of females and males in the COVID-exposed dose 1 group was similar (1075 vs 1255.5). However, as shown in **Figure 3**, these differences were not statistically significant, although the trend toward a sex difference was more apparent in the COVID-19-naïve dose 1 and dose 2 groups.

**Figure 3.**
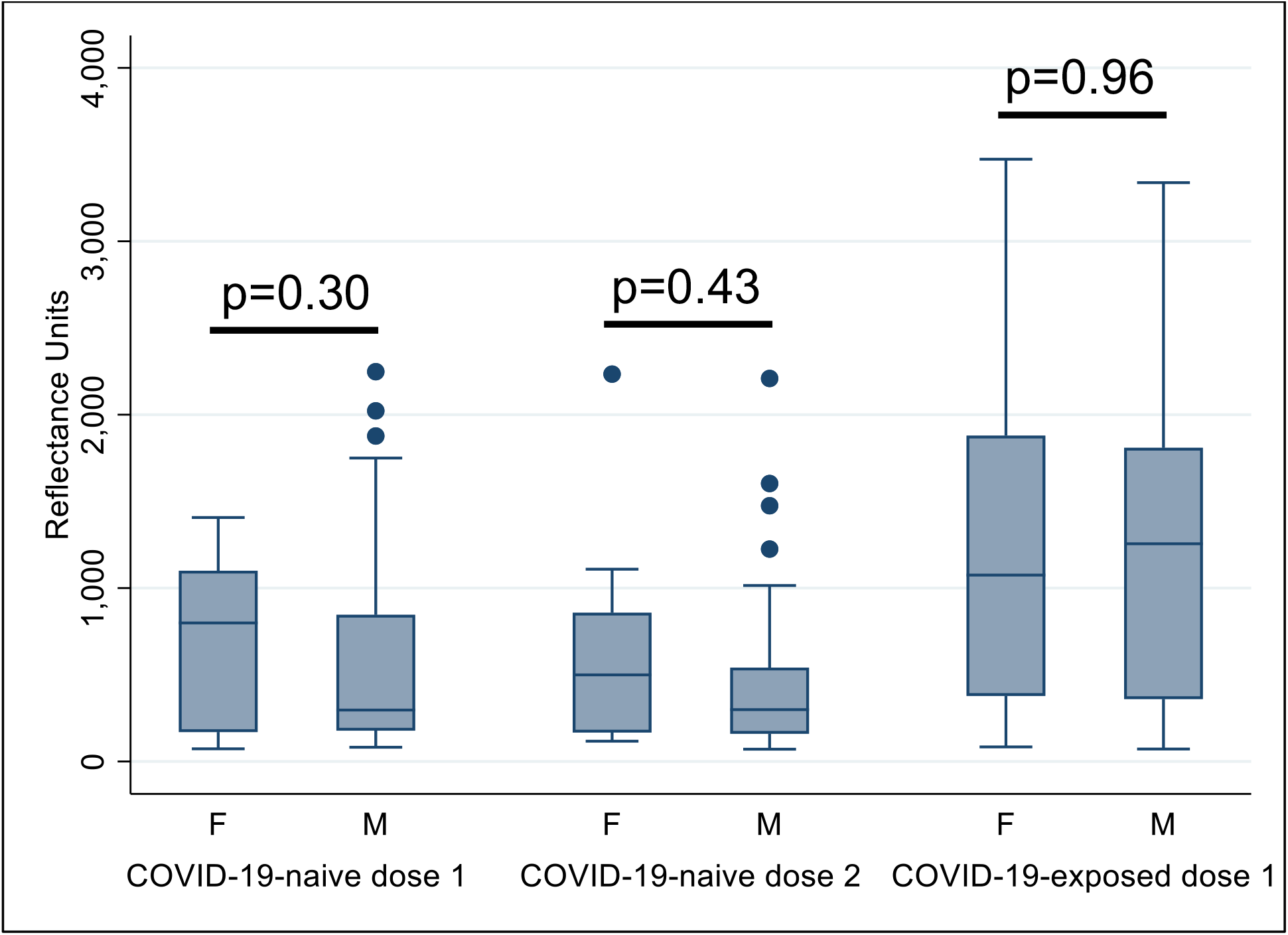
Antibody responses to vaccination in females and males. P values are based on a two-sample Wilcoxon rank-sum (Mann-Whitney) test.

## DISCUSSION

In this study, we describe the performance of a novel, rapid, oral-fluid-based diagnostic device (CovAb™) for determination of infection or vaccine-induced anti-SARS-CoV-2 antibodies. We evaluated test performance in two distinct COVID-19 patient populations, one in the US and one in India. Our findings demonstrate that the CovAb™ test exhibits similar performance in both geographical populations and generates results in accordance with the commercial blood-based serology test. We also investigated antibody responses in both COVID-naïve and COVID-exposed (nursing home staff) groups as well as responses to first and second doses of vaccine in the COVID-naïve group. We found that there was a wide range of response after both the first and second dose of the Oxford-AstraZeneca vaccine, with significant numbers of vaccinated individuals lacking a detectable response, even following a second dose, although the antibody response was greater following the second dose. A more robust serology response to the second dose has also been recently reported for mRNA vaccines (36). We also found that antibody responses were more prevalent and robust in a group with prior exposure to COVID-19 patients. An enhanced serology response to mRNA vaccination in previously infected individuals has been reported (37,38), but this appears to be the first report of enhanced response in asymptomatic COVID-exposed individuals.

The CovAb™ test employs an oral fluid sample enriched in GCF vs saliva. While SARS-CoV-2 antibodies have been reported in saliva following infection, those saliva samples presumably included GCF, since most saliva collection protocols for diagnostic testing do not obtain pure saliva. Our preliminary data comparing GCF vs saliva-enriched oral fluid indicate that GCF is the major source of the SARS-CoV-2 antibodies detected with the CovAb™ test, consistent with its characterization as a plasma transudate and which is responsible for oral fluid IgG, IgM, and monomeric IgA (39).

The CovAb™ test, by virtue of its POC format, noninvasive sample collection, and rapid time frame, should be of general use for detection of previous (within several months) infection or initial adaptive immune response to vaccination. Verification of effective post-vaccination antibody response will be important to determine the need for use of a different vaccine, or booster of the same vaccine, and to predict the likelihood of subsequent memory B cell development responsible for long-term immunity (40,41).

## Data Availability

All data referred to in the manuscript are included in the data shown.

